# False Negative Mitigation in Group Testing for COVID-19 Screening

**DOI:** 10.1101/2020.07.31.20154070

**Authors:** A. R. Alizad-Rahvar, S. Vafadar, M. Totonchi, M. Sadeghi

## Abstract

After lifting the COVID-19 lockdown restrictions and opening businesses, screening is essential to prevent the spread of the virus. Group testing could be a promising candidate for screening to save time and resources. However, due to the high false-negative rate (FNR) of the RT-PCR diagnostic test, we should be cautious about using group testing because a group’s false-negative result identifies all the individuals in a group as uninfected. Repeating the test is the best solution to reduce the FNR, and repeats should be integrated with the group-testing method to increase the sensitivity of the test. The simplest way is to replicate the test twice for each group (the 2Rgt method). In this paper, we present a new method for group testing (the groupMix method), which integrates two repeats in the test. Then we introduce the 2-stage sequential version of both the groupMix and the 2Rgt methods. We compare these methods analytically regarding the sensitivity and the average number of tests. The tradeoff between the sensitivity and the average number of tests should be considered when choosing the best method for the screening strategy. We applied the groupMix method to screening 263 people and identified 2 infected individuals by performing 98 tests. This method achieved a 63% saving in the number of tests compared to individual testing. Our experimental results show that in COVID-19 screening, the viral load can be low, and the group size should not be more than 6; otherwise, the FNR increases significantly. A web interface of the groupMix method is publicly available for laboratories to implement this method.

## I. Introduction

**I**n the post-COVID-19 era, most countries are trying to lift their lockdown restrictions. However, an infected person can remain completely asymptomatic and spread the virus. Hence, it is essential to increase screening tests to identify and quarantine the infected person and identify any other person who has been exposed to that individual. Unfortunately, the testing capacity of many countries is not sufficient, and they have to save their capacity for the second or even the third wave of the coronavirus outbreak.

In this situation, the group testing (or pool testing) technique can immediately and dramatically increase worldwide testing capacity by decreasing the required number of individual tests in a population. In this technique, separate samples are mixed together to create a sample pool. After a single diagnostic test has been performed on each group of samples, a negative result indicates that everyone in the group is uninfected; otherwise, at least one of the samples in the group is infected. Then, in conventional group testing [1], [2], an individual test on each sample in the infected group is performed to identify the infected samples. With this method, the number of tests is decreased by not testing the individual samples in an uninfected group.

Although different types of reverse-transcription polymerase chain reaction (RT-PCR) tests are the predominant diagnostic methods for detecting SARS-CoV-2, the accuracy of these methods for COVID-19 is still unknown [3]. False positives are rare for RT-PCR testing when primers and probes are designed appropriately. There are many reports of specificity of 100% for SARS-CoV-2 RT-PCR assays, based on the in vitro cross-reactivity assessment [4], [5], [6]. However, the clinical specificity, which is affected by the contamination of laboratory equipment and reagents or human error, could be less than 100%. Overall, the false positive rate (FPR) of SARS-CoV-2 RT-PCR diagnosis, without human error and contamination, can be considered to be zero.

The major problem in COVID-19 diagnosis with RT-PCR is false negatives. A false-negative rate (FNR) of up to 50% has been reported for RT-PCR-based SARS-CoV-2 diagnostic testing [4], but an FNR of 10-20% is more frequent in the reports [6], [7]. An unsuitable type of sample (e.g., a throat swab rather than a nasal swab [8]) and inadequate or inappropriate specimen collection, storage, and transport are responsible for a large portion of this high FNR. Moreover, the FNR of SARS-CoV-2 RT-PCR diagnosis is affected by the number of days that have elapsed since an individual first became infected. Indeed, the FNR is about 67% and 38% on day 4 and on the day of symptom onset (day 5), respectively. Three days after symptom onset (day 8), the FNR decreases to 20%, and then, it gradually increases again to 66% on day 21 [9].

A false-negative result puts the whole society at risk by falsely indicating that an infected person does not have an infection. Hence, this person, might move around the community and infect others. False negatives in group testing are much riskier than in individual testing. If a group of specimens is infected, each sample in the group can potentially be infected. Consequently, if the test result of this infected pool is a false negative, this result indicates that every person with a specimen in the pool is infection-free. Also, the mixing of specimens in group testing makes an infected specimen become diluted by the uninfected samples. Therefore, if the group size is not determined wisely, the infected specimen becomes undetectable, and the sensitivity of the test is reduced, resulting in false negatives (the dilution effect). Hence, the group testing methods must be made resistant to false-negative results. Unfortunately, the main focus of most of the studies in the field of group testing is only on reducing the average number of tests. Hence, more studies are needed to mitigate the effect of false negatives and increase the sensitivity of the test in group testing. This paper aims to address this need.

In this work, we propose a new method of group testing, called the groupMix method, for mitigating the false negatives. We propose 1-stage and 2-stage sequential versions of this method. In [10], the authors propose a method for false-negative mitigation of SARS-CoV-2 group testing. We propose a 2-stage version of this method and compare the sensitivity and the average number of tests of 1-stage and 2-stage versions of this method with those of the groupMix method. Then we present our experimental test results for COVID-19 screening by using the groupMix method. Finally, we introduce our web interface, which will help laboratories to implement the groupMix method.

## II. Group Testing Methods

In the case of individual testing, the best practice to mitigate the effect of false negatives is to repeat the test [11]. If the false-positive results are ignored, a person can be diagnosed as positive if either test is positive. Assume that there is no human error and contamination resulting in false positive, and the FNR of the RT-PCR test is 20% and the test errors are independent. In this case, if we repeat the test for an individual twice independently by getting two samples, the chance of obtaining two false-negative results drops to 4%. Consequently, the sensitivity of the test increases from 80% (for a single test) to 96% (for two tests). Inspired by this easy method of false-negative mitigation for individual testing, this test repetition could be integrated into the group-testing procedure to mitigate the effect of false negatives in the detection of infected groups. For instance, an adaptive screening procedure is introduced in [12] so that the negative groups of each stage are re-tested. If both tests of a group are negative, this group is considered as a negative group. Recently, a group testing method has been proposed for SARS-CoV-2 in [10] for false-negative mitigation. Here, we call this method 2-replicate group testing, denoted by 2Rgt. We introduce this method in the next section, and then, we propose a 2-stage sequential version of it to reduce its required number of tests. We will compare the results of our proposed method, groupMix, with 2Rgt’s results.

### A. Model and Notation

Assume that the prevalence (prior probability) of the disease in the population is *p*. We want to test *N* independent specimens by using group testing. The group size, i.e., the number of specimens in each pool, is *n*. The optimum group size, *n*_*opt*_, minimizes the average number of tests, 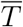. The FNR of the diagnostic method for a single test is denoted by *f*_*N*_, and the FPR is negligible. It is assumed that testing the pooled sample does not change *f*_*N*_. In other words, the group size *n* is determined wisely to prevent the dilution effect and a drop in the sensitivity.

Denoting a group of samples by *G, G* = 0 means that all samples in *G* are uninfected. In contrast, *G* = 1 implies that *G* is a positive group; i.e., it has at least one infected specimen. By observing the results of group tests in different methods, we identify a group as infected or uninfected, denoted by *Ĝ* = 1 and *Ĝ* = 0, respectively. Here, we define two types of sensitivity. The *sample-level sensitivity, S*_*s*_, refers to the probability of detecting an infected sample as positive. On the other hand, the *group-level sensitivity, S*_*g*_, refers to the probability of positive detection of an infected group; i.e., *P* (*Ĝ* = 1 |*G* = 1). To be able to detect an infected sample, its corresponding pool should be diagnosed positive first, and then, its individual test should also be positive. Therefore, *S*_*s*_ = *S*_*g*_(1 −*f*_*N*_). In comparing different methods of group testing, we compare *S*_*g*_ values to measure the accuracy of the methods.

### B. groupMix Method

The schematic diagram of the groupMix method is depicted in Fig. 1. This method has the following steps.

**Fig. 1.**
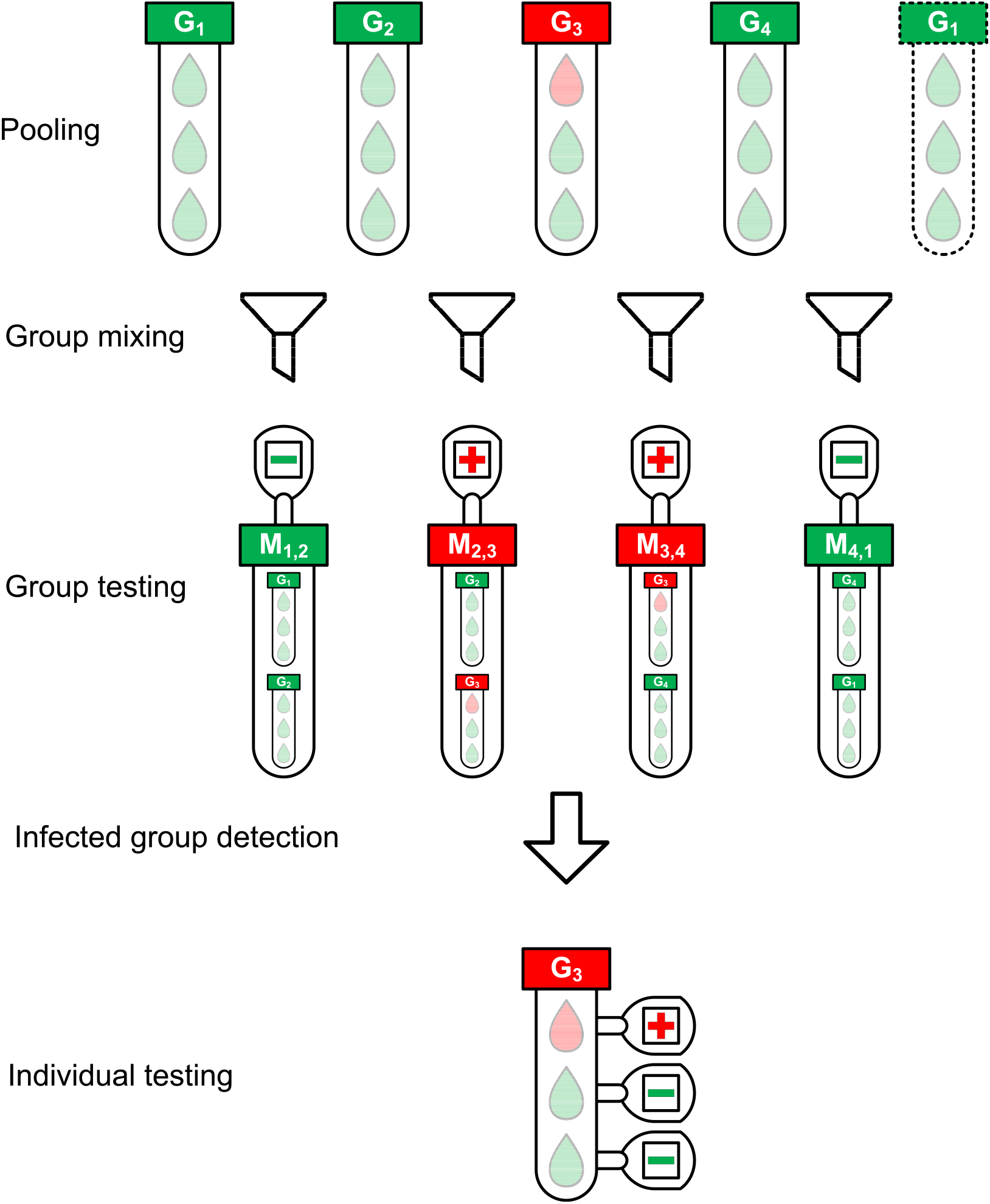
The schematic diagram of the groupMix method for non-conservative infected group detection.

1. **Pooling:** We make groups of samples with the group size of *n*. Therefore, we have *m* = *N/n* groups. We label each group as *G*_*i*_ where 1 ⩽ *i ⩽ m*, and we call them *primary groups*.
2. **Group mixing:** Each primary group *G*_*i*_ (2 ⩽*i ⩽m* − 1) is mixed with *G*_*i*−1_ and *G*_*i*+1_ separately, giving the mixed groups *M*_*i*−1,*i*_ and *M*_*i,i*+1_, respectively, a group size of 2*n*. In the case of the first (*G*_1_) and last (*G*_*m*_) groups, we mix them together and make *M*_*m*,1_. With this form of mixing, each group exists in two mixed groups. Hence, each group will be tested twice by testing the mixed groups.
3. **Group Testing:** Each mixed group will be tested to identify the infected mixed groups. This form of group mixing results in *m* mixed groups, which equals the number of the primary groups *G*_*i*_. Therefore, in this step, we need *m* tests.
4. **Infected group detection:** In this step, we detect the infected primary groups from the test results of the mixed groups. Different approaches can be applied for this purpose, based on the level of compromising the false negatives and the tradeoff between sensitivity and the number of tests. Assume that we want to detect the infected group in Fig. 1. Here, *M*_2,3_ and *M*_3,4_ are positive. We can have the following detection approaches.

- **Conservative group detection:** In a conservative approach, *M*_2,3_ = 1 indicates that both *Ĝ*_2_ and *Ĝ*_3_ should be considered as infected because both can cause a positive result for *M*_2,3_. However, *Ĝ*_2_ = 1 should also make the test result for *M*_1,2_ positive. Here, we assume pessimistically that the observed value of *M*_1,2_ = 0 is a false-negative result. Similarly, *M*_3,4_ = 1 results in *Ĝ*_3_ = 1 and *Ĝ*_4_ = 1. Hence, with this approach, the primary groups *Ĝ*_2_, *Ĝ*_3_, and *Ĝ*_4_ should be detected as positive in Fig. 1. Here, *Ĝ*_2_ = 1 and *Ĝ*_4_ = 1 are false positives due to our detection algorithm. Consequently, this method makes algorithmic false positives, increasing the number of tests. However, this approach results in 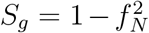. In summary, in conservative group detection, we consider *Ĝ*_*i*_ = 1 and *Ĝ*_*i*+1_ = 1 provided that *M*_*i,i*+1_ = 1.
- **Non-conservative group detection:** To reduce the number of algorithmic false positives, and consequently, to decrease the number of tests in conservative group detection, we should be tolerant of false negatives. To achieve this goal, we can detect only more probable infected groups, with the cost of less sensitivity. In this way, both *M*_2,3_ and *M*_3,4_ in Fig. 1 are positive, most likely because *G*_3_ = 1. Although these positive results can also occur if *G*_2_ = 1 and *G*_4_ = 1, the lack of *M*_1,2_ = 1 and *M*_4,1_ = 1 makes it unlikely that *G*_2_ and *G*_4_ will both equal 1. In this approach, if, for example, *G*_2_ is truly positive and *M*_1,2_ = 0 is a false-negative result, we will miss *Ĝ*_2_ = 1, resulting in less sensitivity. We will propose a 2-stage sequential version of this method to resolve this problem and increase the sensitivity.

If there is not any false-negative result, we should always see a pair of positive mixed groups for an infected group *G*_*i*_, i.e., *M*_*i*−1,*i*_ and *M*_*i,i*+1_. Therefore, provided that we observe only a single positive mixed group, i.e., *M*_*i*−1,*i*_ = 0, *M*_*i,i*+1_ = 1, and *M*_*i*+1,*i*+2_ = 0, then the other mixed group of the pair is not diagnosed positive because of the false-negative result. Hence, a single positive mixed group *M*_*i,i*+1_ indicates that both *G*_*i*_ and *G*_*i*+1_ can potentially be infected. Consequently, we consider both groups positive to test all of their specimens individually in the next step.

In summary, we have the following rules in non-conservative group detection.

1. Provided that the test result of both *M*_*i*−1,*i*_ and *M*_*i,i*+1_ are positive, *Ĝ*_*i*_ is considered as a positive group; otherwise, it is negative.
2. If we have a single positive mixed group, e.g., *M*_*i,i*+1_, both *Ĝ*_*i*_ and *Ĝ*_*i*+1_ are considered as infected.

**5) Individual testing:** The specimens of all primary groups that are detected positive (*Ĝ*_*i*_ = 1) are tested individually.

In Fig. 1, there are *N* = 12 samples with one infected sample. The pooling of samples is performed with a group size of *n* = 3. Hence, there are *m* = 4 primary groups in which *G*_3_ is infected. Fig. 1 depicts the non-conservative group detection. We need 4 tests in the group-testing step and 3 individual tests in the last step, or a total of 7 tests. Using the conservative group detection, we need 4 tests in the group-testing step and 9 more individual tests, or a total of 13 tests. We can see the effect of the algorithmic false positives in increasing the number of tests.

### C. Sequential groupMix Method

We propose a 2-stage sequential version of the non-conservative groupMix method, denoted by 2S-groupMix, to increase its sensitivity. As explained in Sec. II-B, some cases are undetected by the non-conservative group detection. For instance, assume that *G*_1_ = 0, *G*_2_ = 1, *G*_3_ = 0, and *G*_4_ = 1. In this case, if we observe *M*_1,2_ = 0 (a false negative), *M*_2,3_ = 1, *M*_3,4_ = 1, and *M*_4,1_ = 1, the non-conservative method detects *Ĝ*_3_ = 1 and *Ĝ*_4_ = 1. Here, *Ĝ*_2_ = 1 is not detected because of the false-negative result. Since *Ĝ*_3_ = 1 is an algorithmic false positive, the individual test of its specimens will not show any infected sample. This observation indicates that *M*_2,3_ = 1 could be due to *G*_2_ = 1, and implicitly shows that *M*_1,2_ = 0 is a false negative. Therefore, in the second stage, *Ĝ*_2_ is considered infected, and its specimens are tested individually. In this way, the sensitivity of the non-conservative groupMix method is increased by performing the second stage. In summary, the 2S-groupMix method is as follows.

1. Perform the groupMix method by using the non-conservative group detection.
2. Assume that the test of a mixed group is positive in the previous stage (e.g., *M*_*i,i*+1_ = 1), and that only one of its primary groups is detected as a positive group (for example, *Ĝ*_*i*_ = 0 and *Ĝ*_*i*+1_ = 1). Provided that the individual tests on the samples of the detected group (i.e., *G*_*i*+1_) are all negative, perform the individual test on the specimens of the other undetected primary group (i.e., *G*_*i*_).

Since this approach is a sequential method, we need to spend more time to increase the sensitivity of the test. In other words, there is a tradeoff between the test time and the sensitivity.

### D. 2-replicate group test

In 2-replicate group test (2Rgt) method, each group is tested twice. Provided that the result of at least one of the tests is positive, this group is diagnosed as a positive group. In this case, 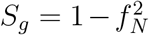.

Note that the 2Rgt method has no algorithmic false positive.

In the case of Fig. 1, we need 11 tests if we use the 2Rgt method, while 7 and 13 tests are required if we use the non-conservative and the conservative groupMix method, respectively.

### E. Sequential 2-replicate group test

Here, we present a 2-stage sequential version of the 2Rgt method, denoted by 2S-2Rgt, to reduce the number of tests, while the sensitivity of the test, i.e., 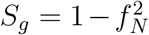, remains the same. The drawback of the 2Rgt method is that it uses two tests for each group and increases the required number of tests.

Since observing only one positive result out of two tests is enough to consider a group as infected, we can separate these two tests on each group and avoid the second test for positive groups of the first stage. The summary of this method is as follows.

1. Test all groups once and identify the positive groups as infected.
2. Perform the second test only on the negative groups of the first stage and identify the positive groups.
3. Individually test the specimens of the positive groups of stages 1 and 2.

In this way, we reduce the number of tests by saving the second test of the infected groups identified in the first stage.

## III. Analytical Results

In this section, we present the results of the analytical analysis of the different group-testing methods regarding *S*_*g*_, and the average number of tests per sample, 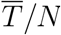. The proof for the analyses is available in Appendix A. All analytical results are verified by simulation. In the following results, it is assumed that the FNR of the diagnostic test is 10%.

### A. The effect of group size on 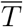 and S_g_

First, we consider the effect of the group size on 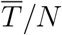 and *S*_*g*_ in Fig. 2 for FNR = 10% and *p* = 2%. This figure has two different y-axes. The right axis shows *S*_*g*_ for the non-conservative (NC), conservative (C), and 2-stage sequential (2S) groupMix methods, the 2Rgt and 2S-2Rgt methods, and the conventional method. The left axis shows 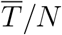 for the NC-groupMix and the 2S-groupMix methods. This figure indicates that for these specific values of FNR and prevalence, *n*_*opt*_, which minimizes the average number of tests, is 7.

**Fig. 2.**
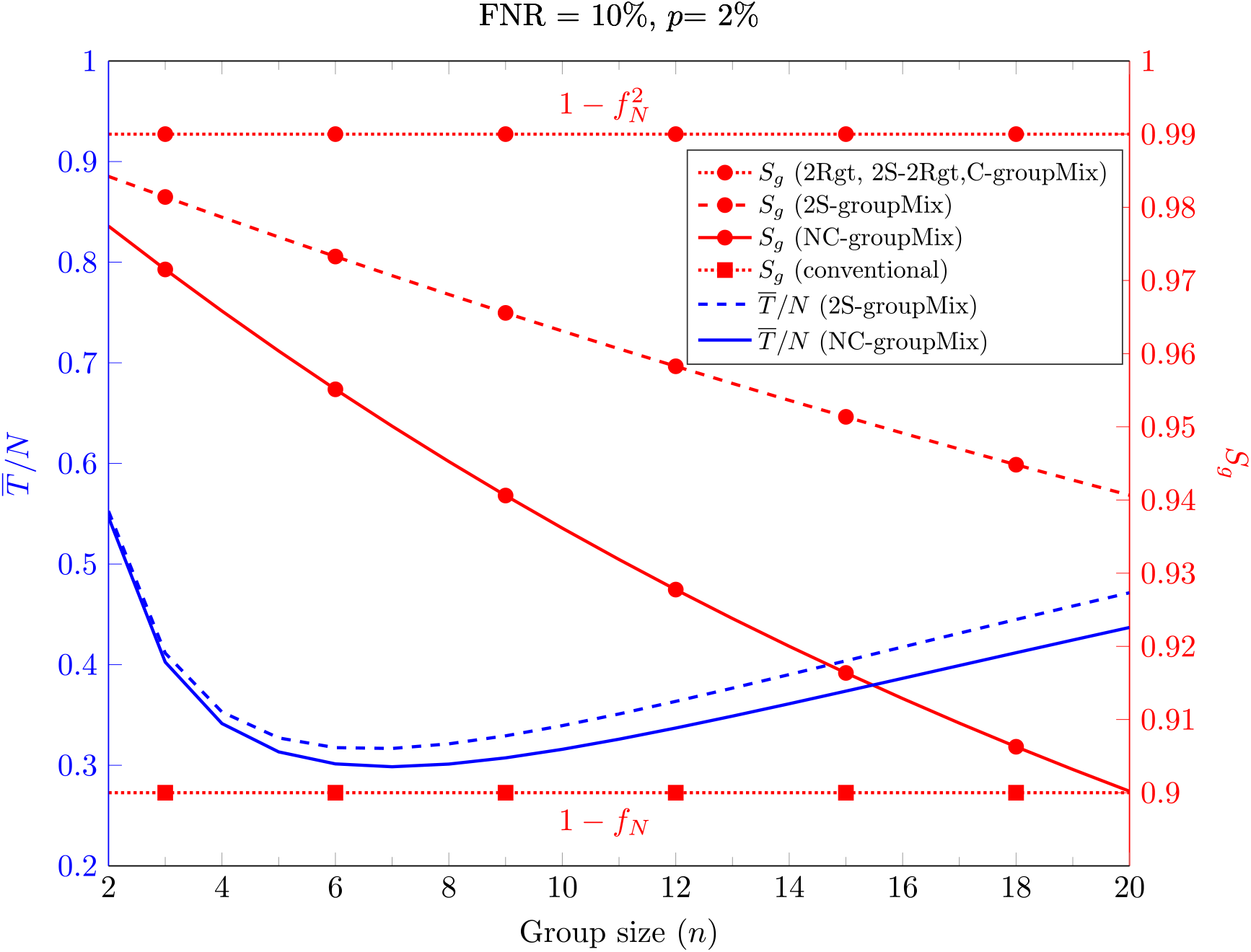
The average number of tests per sample (left y-axis, blue curves) and group-level sensitivity *Sg* (right y-axis, red curves) of different methods against the group size.

Moreover, Fig. 2 shows that *S*_*g*_ of both the C-groupMix and 2Rgt methods is independent of *n*, equal to 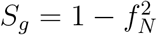, which is 0.99 for *f*_*N*_ = 0.1. Similarly, the conventional group-test method has the fixed group-level sensitivity *S*_*g*_ = 1 −*f*_*N*_ over *n*. However, *S*_*g*_ in NC- and 2S-groupMix methods varies with *n*. In other words, with smaller value of *n*, a larger value of *S*_*g*_ is achieved. In this figure, it is obvious that there is a tradeoff between *S*_*g*_ and 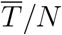 for *n ⩽*7.

If the primary concern in group testing is only to reduce the average number of tests 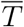, then the group size should minimize 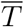. However, in practice, using the optimum value of *n* may not be possible in the presence of problems like the dilution effect. For example, the study in [13] shows that for detecting SARS-CoV-2 with the standard kits and protocols, a single infected sample can be detected in pools of up to 32 samples, with an estimated FNR of 10%. This finding could be valid for the specimen of an active symptomatic patient. However, in COVID-19 screening, we usually deal with asymptomatically-infected or pre-symptomatic individuals or people with very mild or atypical symptoms. These cases may not have a high viral load and detectable amounts of the virus in their specimen.

Consequently, in the screening scenario, the maximum group size could be much less than 32. Our experiments show that a group size of 6 is a good choice for group testing in COVID-19 screening; otherwise, the viral load in the pool becomes very low, and the FNR increases dramatically. A thorough investigation is required to determine the maximum group size for COVID-19 screening rather than for diagnostic testing for detecting active symptomatic patients.

### B. Optimum group size

Figure 3(A) depicts *n*_*opt*_ against the prevalence percentage *p* for FNR = 10%. In the case of the groupMix methods, *n*_*opt*_ refers to the optimum group size of the primary group, i.e., *G*_*i*_. Hence, the optimum size of the mixed group tested by the diagnostic method is 2*n*_*opt*_. Theoretically, the smaller values of *p* allow us to have larger groups of specimens. However, as we discussed above, the use of larger groups can reduce the sensitivity of the test.

**Fig. 3.**
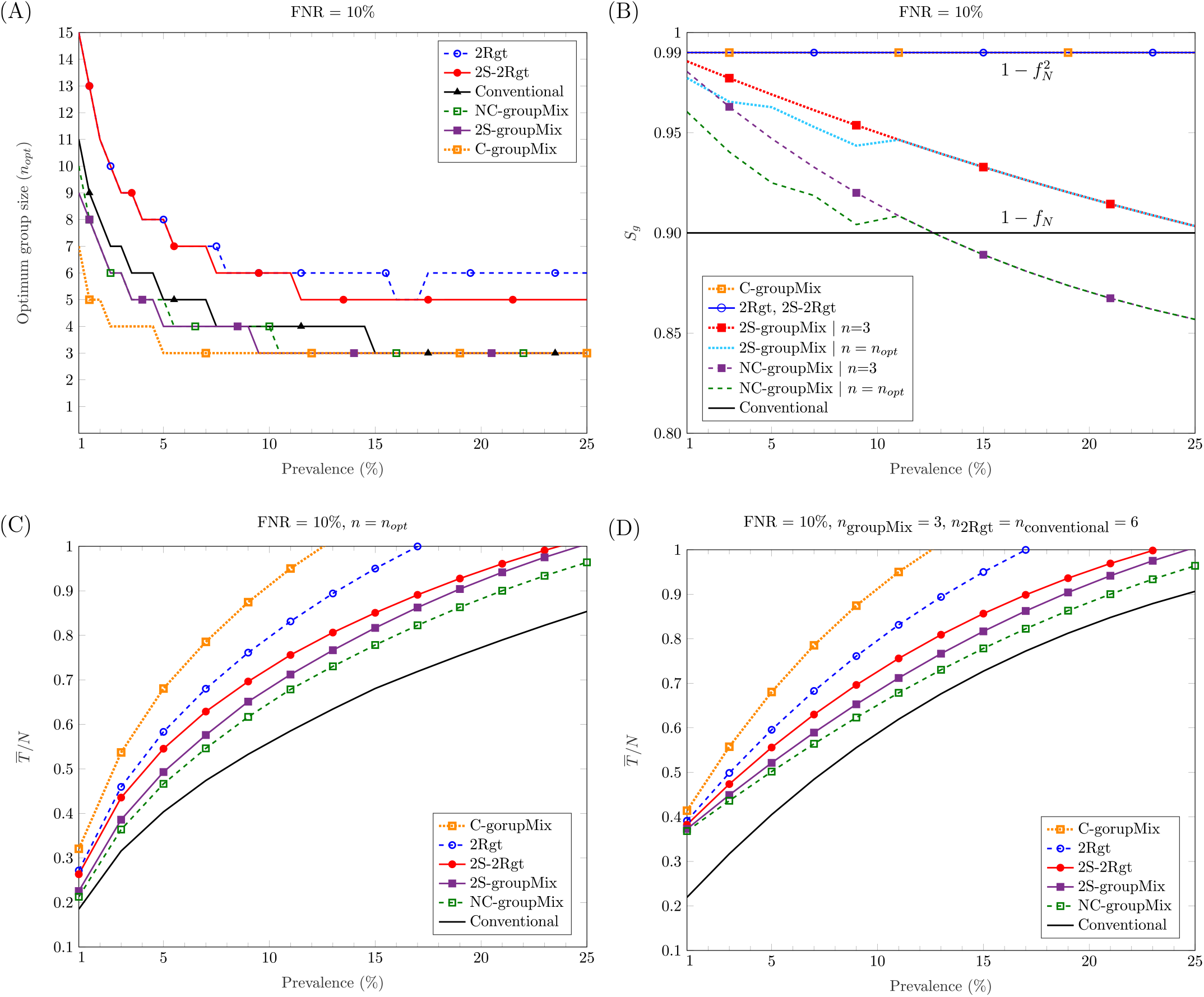
Comparing different methods against the percentage of prevalence: (A) the optimum group size *nopt* to minimize the average number of tests; (B) the group-level sensitivity *Sg* of the methods; (C) the average number of tests per sample for *n* = *nopt*; (D) the average number of tests per sample for the group (or the mixed group in the groupMix method) size of 6.

### C. Group-level sensitivity

Figure 3(B) shows *S*_*g*_ versus *p* for FNR= 10%. Overall, *S*_*g*_ of the C-groupMix, 2Rgt, and 2S-2Rgt methods does not vary with *p* and has the fixed value of 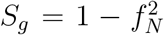. Similarly, the conventional group-test method has the *p*-independent sensitivity of *S*_*g*_ = 1 −*f*_*N*_. However, when we utilize the non-conservative infected group detection in the groupMix method, i.e., the NC- and 2S-groupMix methods, *S*_*g*_ is a decreasing function of *p*. In non-conservative group detection, the sensitivity is compromised by accepting more false negatives to reduce the number of algorithmic false positives. The undetected cases in the non-conservative method occur more in higher values of *p*, resulting in less sensitivity. Even after a specific value of *p*, e.g., *p >* 0.13 for the NC-groupMix method and the FNR of 10%, *S*_*g*_ becomes less than that of the conventional method. Therefore, the usage of the NC-groupMix method is acceptable for small values of *p*, say, less than 5%. Moreover, Fig. 3(B) depicts that the 2S-groupMix method improves *S*_*g*_ significantly compared to the NC-groupMix method.

As we saw in Fig. 2, *S*_*g*_ of the C-groupMix, the 2Rgt, and the conventional methods does not vary with *n*. On the contrary, *S*_*g*_ of the NC-groupMix and the 2S-groupMix methods depends on *n*. For these two methods, we see *S*_*g*_ in Fig. 3(B) for two cases of *n* = 3 (i.e., a mixed group size of 6) and *n* = *n*_*opt*_. For FNR = 10%, *n*_*opt*_ equals 3 for *p* greater than about 10%, resulting in the same *S*_*g*_ for two different cases of group size. For smaller values of prevalence, *n*_*opt*_*>*3. Since *S*_*g*_ of the NC-groupMix and the 2S-groupMix methods is a decreasing function of *n* (Fig. 2), the group testing with *n* = *n*_*opt*_ is less sensitive than *n* = 3 for *p <* 10%.

### D. Average number of tests

Figure 3(C) depicts the average number of tests per sample, 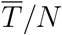, against the prevalence percentage *p* for *n* = *n*_*opt*_. As discussed earlier, in a COVID-19 screening scenario, the group size should not be more than 6 to prevent the dilution effect. Hence, Fig. 3(D) shows 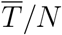 of different methods versus *p* for *n*_groupMix_ = 3 (the size of the mixed group is 6), *n*_2Rgt_ = 6, and *n*_conventional_ = 6. As these figures show, all methods proposed for false-negative mitigation require more tests compared to the conventional method.

Among the 1-stage methods, the NC-groupMix method needs the least average number of tests. However, as mentioned before, its sensitivity reduces with *p*, and after a specific value of *p*, its sensitivity becomes even worse than that of the conventional group test method. Therefore, it is reasonable to use the NC-groupMix method for small values of *p*, say *p <* 5%, to have both high sensitivity and less number of tests. In a comparison between the 1-stage C-groupMix and the 1-stage 2Rgt methods, which both have the same sensitivity of 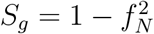, the C-groupMix method needs more tests due to algorithmic false positives. Therefore, if the goal is to reach 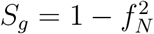 by using a 1-stage method, the 2Rgt method is preferred.

In the case of the 2-stage sequential methods, the average number of tests in the 2S-2Rgt method is less than that of the 2Rgt method. This reduction in the number of tests occurs because, in the 2S-2Rgt method, we perform the second test only for the negative groups in the first stage, rather than using two tests for each group in 2Rgt. In contrast, in groupMix methods using the non-conservative infected group detection, the 2S-groupMix needs more tests compared to the 1-stage NC-groupMix method. This increase in the number of tests occurs because the sequential version performs the full NC-groupMix test in the first stage, followed by the second stage.

In a comparison between the 2S-groupMix and the 2S-2Rgt methods, the former needs fewer tests than the latter. However, the sensitivity of the 2S-groupMix method is less than that of the 2S-2Rgt, and reduces with an increase of *p*. Therefore, there is a tradeoff between *S*_*g*_ and 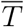 in decisions about choosing a 2-stage sequential group-test method for false-negative mitigation.

Considering the maximum group size (or the size of the mixed group in the groupMix method) of 6, compared to individual testing at 1% prevalence, we save 60% at the FNR of 10% for all proposed methods to mitigate the false negatives. For the optimum group size, this saving is between 70% to 80% at 1% prevalence, but it may result in less sensitive tests.

Overall, in choosing the best group-testing method to mitigate the false-negative results, we should determine the desired level of sensitivity and the amount of saving in the number of the tests. Moreover, the expected time of the test determines whether the sequential method should be used.

## IV. Experimental Methods

We performed COVID-19 screening in the Institute for Research in Fundamental Sciences (IPM), Tehran, Iran. A total of 263 individuals participated in the screening on five different days. These groups consisted of 78, 30, 31, 66, and 58 individuals, respectively. Swabs from the throat were collected and sent to the Clinical Genetic Laboratory at the Royan Institute, Tehran, Iran. Samples were collected between June 15-30, 2020.

The samples were pooled into the primary groups with a size of 3 prior to RNA extraction. Then, the primary groups were mixed according to the groupMix method’s procedure, resulting in mixed groups of size 6.

A volume of 200 µL of the sample was mixed with 600 µL Lysis buffer, and RNA was extracted by using the Norgen Cell-Free RNA Purification Kit (Cat. No: 56300). 50 µL of Elution buffer was used in the extraction procedure.

The Novel Coronavirus (2019-nCoV) Real-Time Multiplex kit (Liferiver, Cat. No: ZJ0009) was utilized for real-time RT-PCR diagnosis. This kit detects the presence of SARS-CoV-2 RNA by detecting the three genes N, OFR1ab, and E. Reactions were heated to 45°C for 10 minutes (1 cycle) for reverse transcription and denatured in 95°C for 90 seconds (1 cycle). Then, 45 cycles of amplification were carried out in 95°C for 15 seconds and 58°C for 30 seconds. Fluorescence was measured at 58°C. The 1-stage NC-groupMix method was performed to detect the infected specimens.

This study was approved by the ethical committee of the Royan institute with a waiver of informed consent due to de-identified nature of the data.

## V. Experimental Results

On the second, fourth, and fifth days, all groups with 30, 66, and 58 individuals were negative. For these days, we performed 10, 22, and 20 tests, respectively. Therefore, we saved 67%, 67%, and 66% in the number of tests, compared to individual testing.

In the first group, consisting of 78 individuals, we had 26 primary and mixed groups. With the NC-groupMix method, only the mixed group *M*_20,21_ was positive. Since we should always have a pair of positive mixed groups, this result indicates that one mixed group is a false negative. Therefore, both primary groups *G*_20_ and *G*_21_ were candidates to be infected. Ultimately, by performing the individual test on these primary groups, one specimen in *G*_20_ was identified as infected. Consequently, we performed 32 tests to detect one infected sample out of 78 samples. Indeed, we had a 59% saving in the number of tests, compared to individual testing.

Regarding the third group consisting of 31 individuals, the mixed groups *M*_4,5_ and *M*_5,6_ were positive. Hence, *G*_5_ was detected as the infected group. The individual test on *G*_5_ revealed the presence of one infected sample in this group. Hence, we detected this infected sample by using 14 tests (i.e., a 55% saving).

In conclusion, 2 positive samples were successfully identified out of 263 by using 98 tests, i.e., a 63% saving in the number of tests compared to individual testing. Since the average prevalence in this population was about 0.8%, the average group-level sensitivity *S*_*g*_ of this screening was very close to 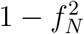.

## VI. Discussion and Conclusion

In this paper, we investigated false-negative mitigation in group testing, focusing on massive testing for COVID-19 screening. Group testing is receiving attention as a strategy that can save time or resources for COVID-19 testing. Indeed, group testing is reasonable for screening since the percentage of infected people is very low, resulting in substantial savings. However, the high rate of false-negative results in RT-PCR-based COVID-19 diagnostic tests has been widely addressed recently as an important public health-related problem. This problem needs more attention in group testing because a false-negative result for a group of potentially infected individuals does not lead to isolating them, and they can quickly spread the virus in their community.

To mitigate the false-negative results and to increase the sensitivity of the group testing, we studied different strategies for repeating the test. By implementing *r* replicates in the test design, we can achieve the maximum group-level sensitivity of 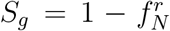, but there is a tradeoff between the sensitivity and the average number of tests; hence, we can reduce the average number of tests by compromising the false negatives. However, this compromising is negligible for the very low percentage of infected samples that usually occurs in screening.

Typically, in group-testing studies, researchers propose different methods for pooling and identifying the infected groups. Then, they find the optimum group size to minimize the average number of tests. However, we explained that in COVID-19 screening, the high viral load might not be available because we are dealing with asymptomatic, pre-symptomatic, or mild-symptomatic people. This fact is crucial in group testing because the dilution effect in the pooling of specimens can cause more false-negative cases. Therefore, the theoretical optimum group size of different methods may yield a severe dilution effect and high FNR in the screening scenario. Our experimental studies showed that the maximum group size in COVID-19 screening should be 6. However, systematic studies are still required to determine the maximum group size for COVID-19 screening. Moreover, by filtering out symptomatic individuals and testing them individually, we can increase the performance and sensitivity of the test. For further studies, we can pool specimens based on the age and medical background of individuals and investigate the effect of these factors on the performance of the group-testing method.

## VII. GroupMix Web Interface

A web interface is developed to help laboratories implement the NC-groupMix method. This website is freely accessible at http://groupmix.ipm.ir. This web interface will be elaborated in Appendix B.

## Data Availability

No data is available.

## Appendix A Analysis

In our analysis, we present *S*_*g*_, and the average number of tests per sample, i.e., 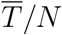, for each method. Then, we explain how to obtain *n*_*opt*_. First, we introduce some notations and basic equations to prove our analysis accordingly.

- #(*x*) ≜ the number of the occurrence of event *x*
- *E*[*X*] ≜ the expected value of a random variable *X*
- *m* ≜ #(groups) = *N/n* (*m* may not equal an integer, however, we do not round it up in our analysis, so that we can calculate the derivatives and simplify the equations. We will round up the all integer size values at the end of our analysis).
- TP_*g*_: the detected positive group is a true positive
- FP_*g*_: the detected positive group is a false positive
- The probability of having a positive group is

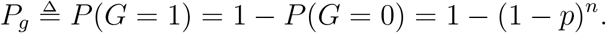
- Group-level sensitivity (true-positive rate TPR_*g*_):

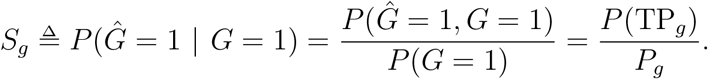

This equation yields *P* (TP_*g*_) = *P*_*g*_*S*_*g*_.
- Group-level FPR:

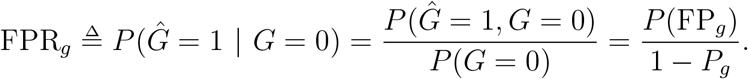

Hence, *P* (FP_*g*_) = (1 − *P*_*g*_) FPR_*g*_.
- 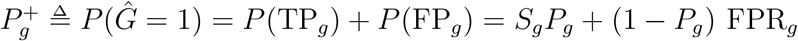.
- 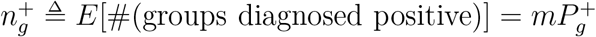.
- Calculation of 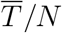 for 1-stage group-test methods:

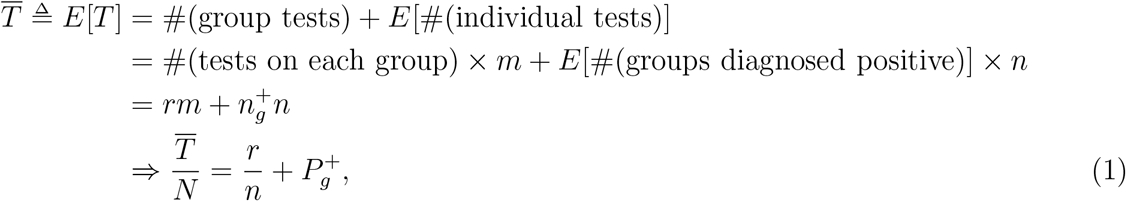

where *r* = 1 for the groupMix and the conventional methods, and *r* = 2 for the 2Rgt method.
- 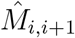: the test result of the mixed group *M*_*i,i*+1_

### A. Analysis of the conventional group-test method

*1) S*_*g*_:

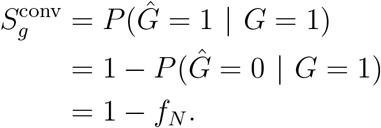

*2)* 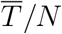: According to Eq. (1) we have

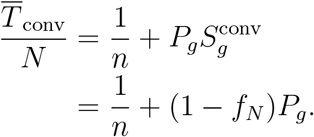

### B. Analysis of the 2Rgt method

Assume that *Ĝ*^(1)^ and *Ĝ*^(2)^ are the results of the first and the second test of *G. Ĝ*^(1)^ and *Ĝ*^(2)^ are dependent because they both depend on whether *G* is infected or not. However, they are conditionally independent given *G*; i.e.,

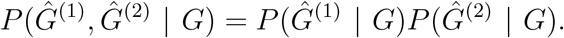

*1) S*_*g*_:

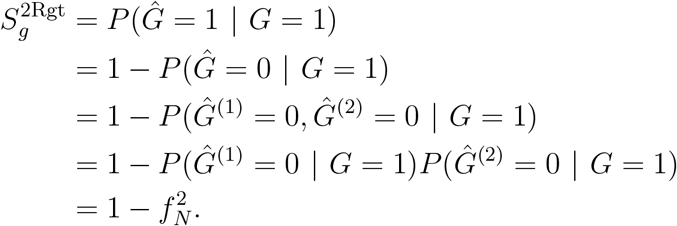

*2)* 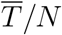: According to Eq. (1) we have

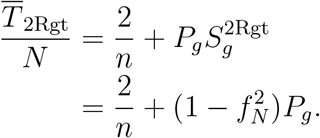

### C. Analysis of the 2S-2Rgt method

*1) S*_*g*_: Assume that *Ĝ*^(1)^ and *Ĝ*^(2)^ are the test results of *G* in the first and the second stage of the sequential 2Rgt (2S-2Rgt) method.

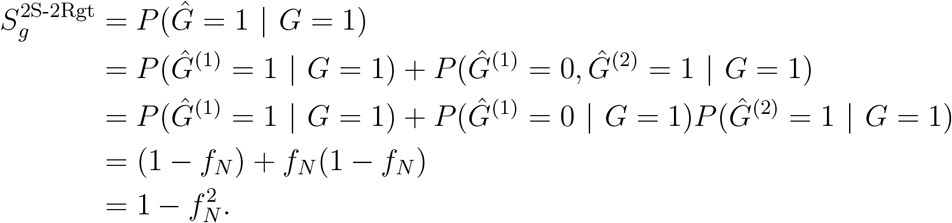

*S*_*g*_ of the 2S-2Rgt is the same as that of the 2Rgt method.

*2)* 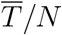:

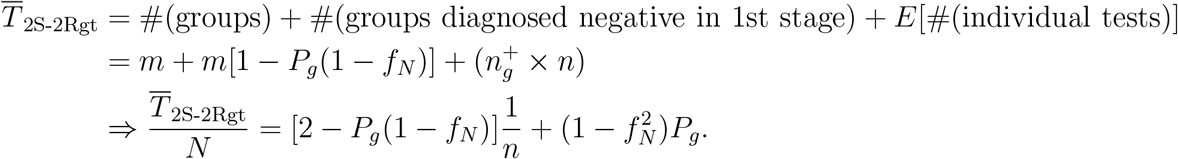

The term 2 − *P*_*g*_(1 *— f*_*N*_) on the right-hand side is less than 2; hence, 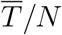 in the 2S-2Rgt method is less than that in the non-adptive 2Rgt method. Indeed, we save on average *NP*_*g*_(1 − *f*_*N*_)*/n* tests with the sequential approach.

### D. Analysis of the C-groupMix method

In the groupMix method, 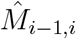 and 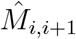 are conditionally independent given *G*_*i*_, i.e.,

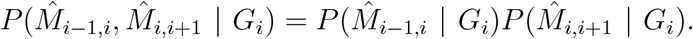

Moreover, we use the following conditional probabilities in the proof of our analyses for the groupMix method.

1. 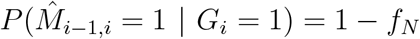
2. 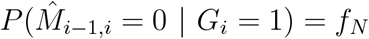
3. 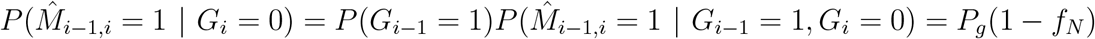
4. 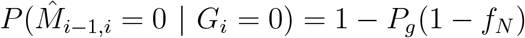

The same equations are valid for 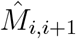.

*1) S*_*g*_:

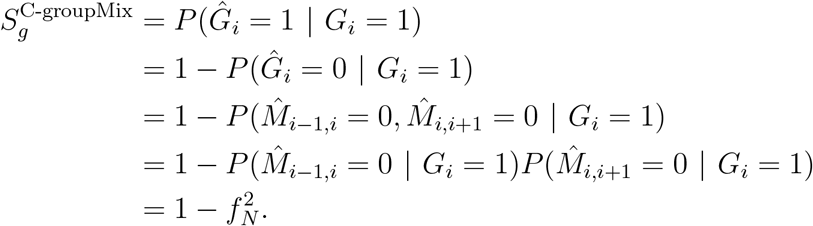

*2)* 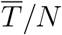: There is an algorithmic false positive in the groupMix method. Hence, we should first obtain the 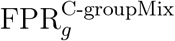 to calculate 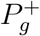 and then calculate 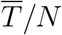.

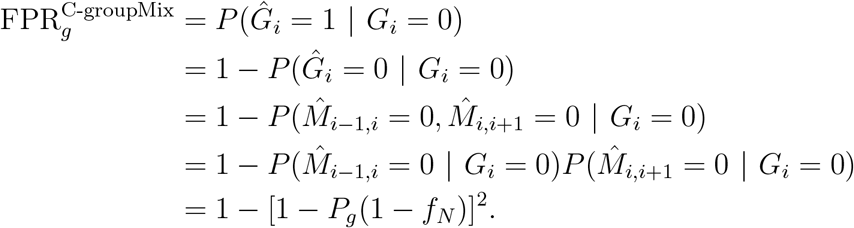

Then, by using Eq. (1) we have

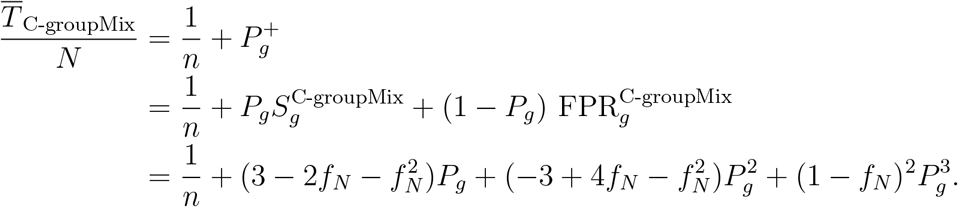

### E. Analysis of the NC-groupMix method

*1) S*_*g*_: In the NC-groupMix method, the primary group *G*_*i*_ is detected positive if the test results of both *M*_*i*−1,*i*_ and *M*_*i,i*+1_ are positive, or if we have a single positive mixed group, i.e., either *M*_*i*−1,*i*_ or *M*_*i,i*+1_. Therefore, we can calculate *S*_*g*_ as follows:

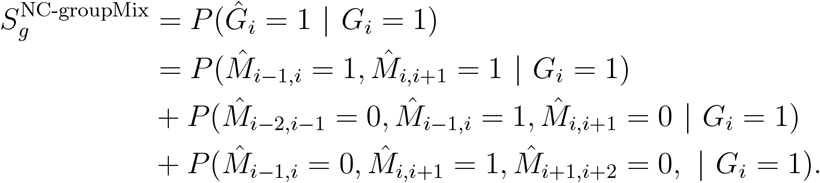

The second and the third probabilities in the above equation are equal. Hence, we have

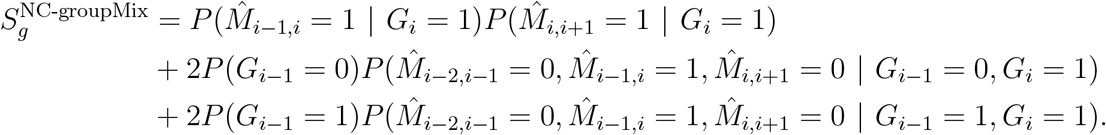

By applying the multiplying property of the conditional independence and using the equations explained at the beginning of Section A-D, we obtain

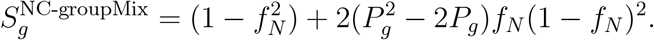

*2)*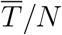: For calculating 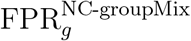, the equations are like the equations for calculating *S*_*g*_, except that the condition is *G*_*i*_ = 0. Ultimately, we obtain

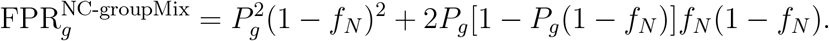

Therefore, we have

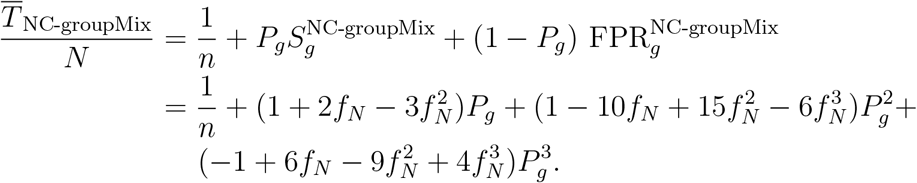

### F. Analysis of the 2S-groupMix method

In the 2S-groupMix method, we first perform the NC-groupMix method, and then, the second stage of the test is done based on the results of the first stage. Therefore, the *S*_*g*_ and 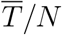 values of the 2S-groupMix method are calculated by using the sum of those values in the NC-groupMix method with the added values obtained by performing the second stage. In the following analyses, the superscript indexes (1) and (2) denote the first and the second stage of the 2S-groupMix method, respectively.

*1) S*_*g*_:

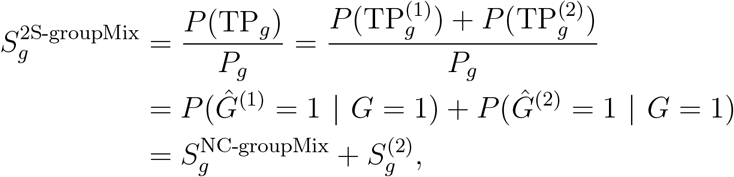

where 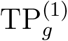 and 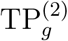mean the true-positive detection of *G* that is detected in the first and second stages, respectively.

In the second stage, the following situations yield a true positive detection of *G*_*i*_ (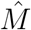 values are the test results of the mixed groups in the first stage):

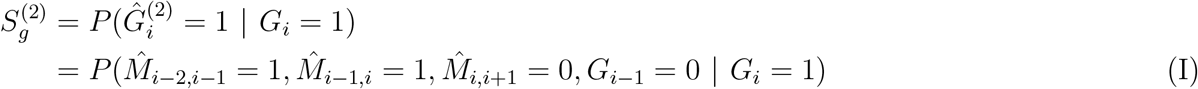

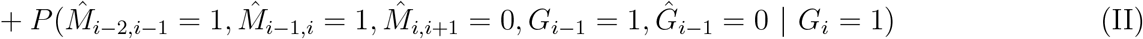

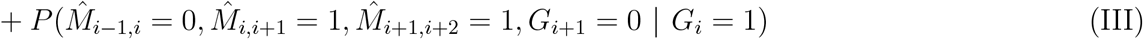

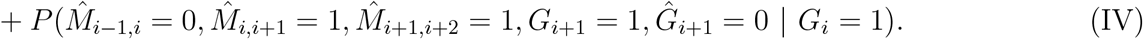

Because of the symmetry, the probabilities (I) and (III), and (II) and (IV) are equal. Therefore, we calculate only the probabilities (I) and (II) separately and then obtain the value of 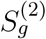.

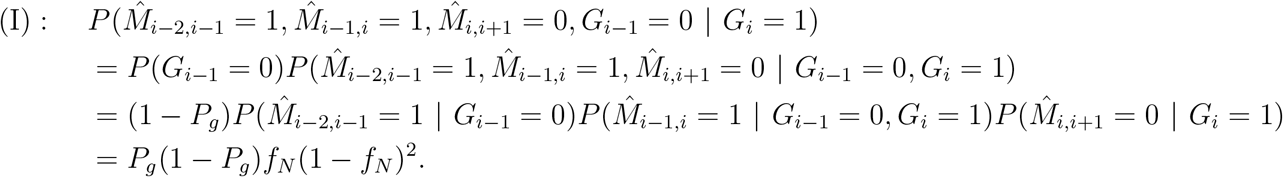

Similarly, we have

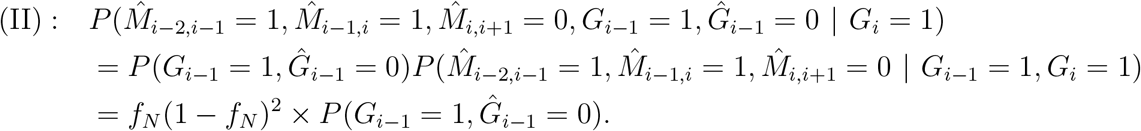

In the above equation, *G*_*i*−1_ = 1 and *Ĝ*_*i*−1_ = 0 means that the primary group *G*_*i*−1_ has at least one infected sample, but the individual test results of the infected samples in the first stage are all false negative. Therefore, *P* (*G*_*i*−1_ = 1, *Ĝ*_*i*−1_ = 0) denoted by Σ (*n, p, f*_*N*_) is as follows:

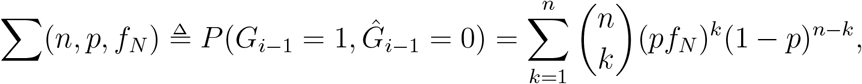

where *pf*_*N*_ is the probability of a sample to be infected and diagnosed negative falsely.

Ultimately, we can obtain *S*_*g*_ of the 2S-groupMix method by using the above results.

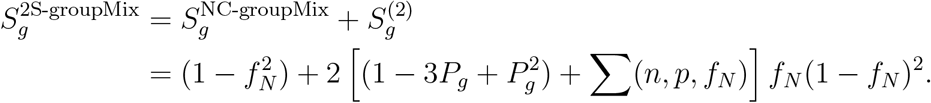

*2)* 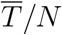: Similarly, we have

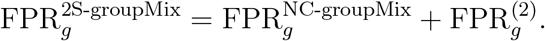

Equations (I) to (IV) used in calculation of 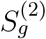 can be used for calculating 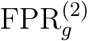 by replacing the condition by *G*_*i*_ = 0. After performing the same calculations, we have

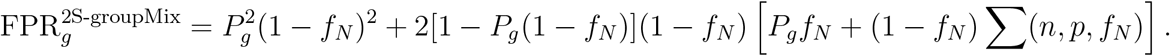

Finally,

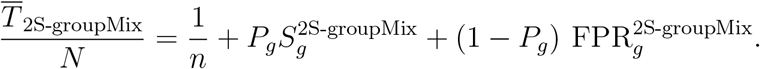

### G. Finding the optimum group size

To find the optimum group size *n*_*opt*_, we should find the minimum value of 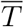 against *n*. Hence, we need to find the solution of 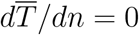. This solution is the same as the solution of 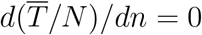.

By using Eq. (1) for 1-stage group-testing methods, we have

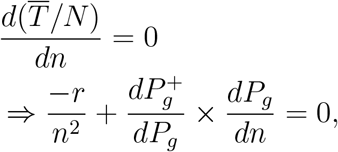

where

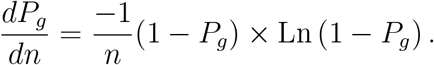

*n*_*opt*_ can be founf by solving the above equation.

In the case of the 2S-groupMix method, 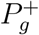 is not only a function of *P*_*g*_, and the derivative of Σ(*n, p, f*_*N*_) does not have a closed-form solution. Therefore, for this method, we find *n*_*opt*_ by using numerical computation. For the 2S-2Rgt method, the calculation of 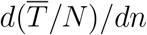 is straightforward when the above formula for *dP*_*g*_*/dn* is used. Ultimately, the value of *n*_*opt*_ obtained by solving the equation 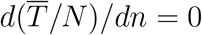 should be rounded to the closest integer number.

## Appendix B GroupMix Web Interface

In this appendix, we explain different parts of the groupMix web interface. In this web, the 1-stage NC-groupMix method is implemented. The following sections in this web interface can be navigated using the left side of the website.

### A. Create Groups

In the “Create Groups” section (Fig. B.1), we should set three input values of the number of specimens, prevalence (%), and FNR (%). Then, the optimum size of the primary groups, i.e.,*n*_*opt*_, the number of groups, and the average number of tests is calculated according to the provided input values. Under these values, the mixed groups and their corresponding specimen numbers are depicted. The group size can be changed to a different value.

For instance, in Fig. B.2, the input values, and the group size are set according to the case of group testing depicted in Fig. 1, where we have 12 samples and four mixed groups. After performing the tests on mixed groups, we can set the test results by clicking on the green icons in the “Test Results” column. The green icon indicates that the mixed group is uninfected, and it turns into a red color, indicating infected, by clicking on the green icon. After determining the infected mixed groups, the index of specimens that should be tested by individual testing, i.e., the samples of infected primary groups detected by the non-conservative group detection method, are listed at the bottom of the page. Finally, this test setup can be saved by clicking on the “Save Test Line” button.

**Fig. B.1.**
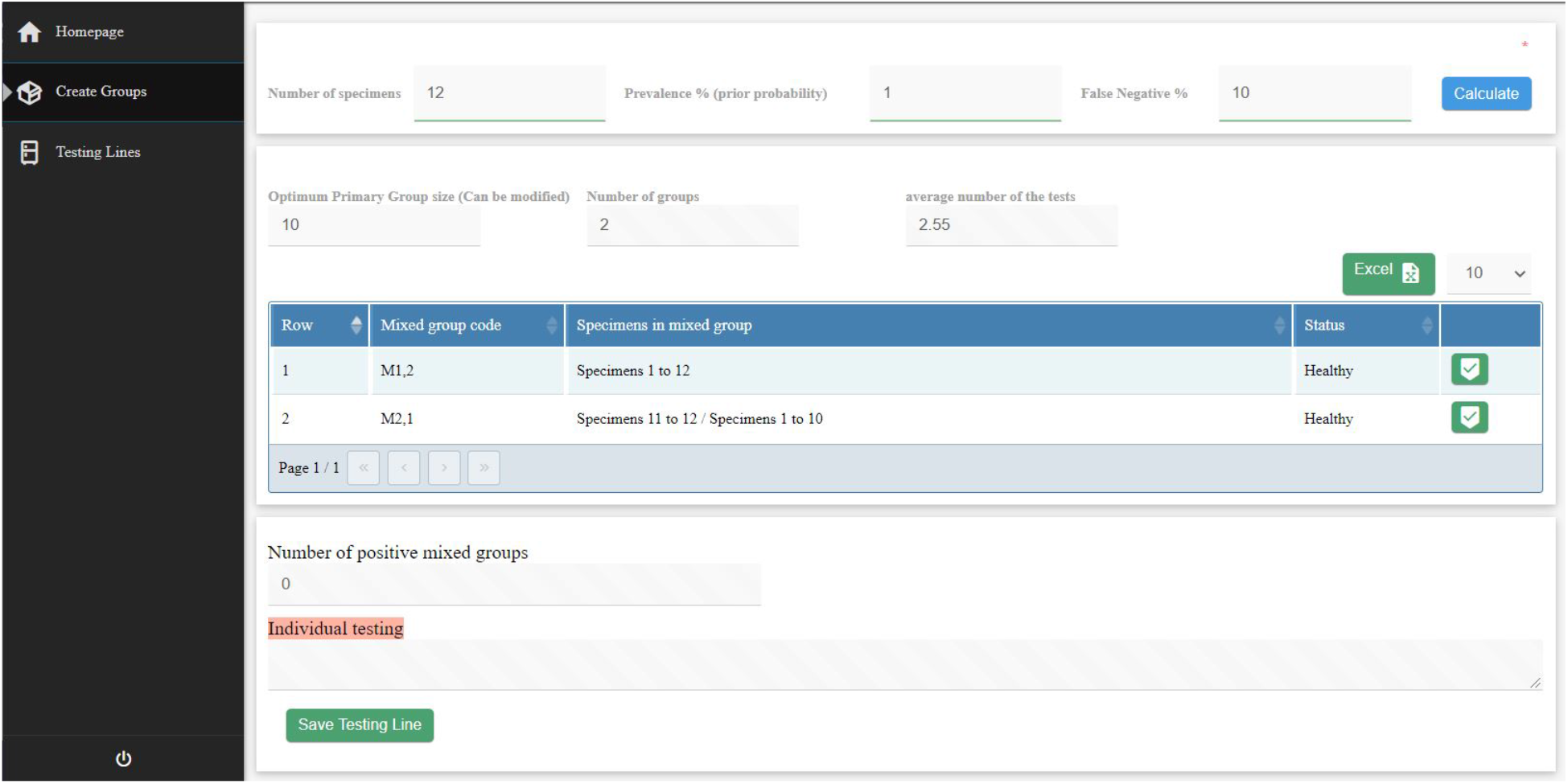
“Create Groups” section in the groupMix web interface.

**Fig. B.2.**
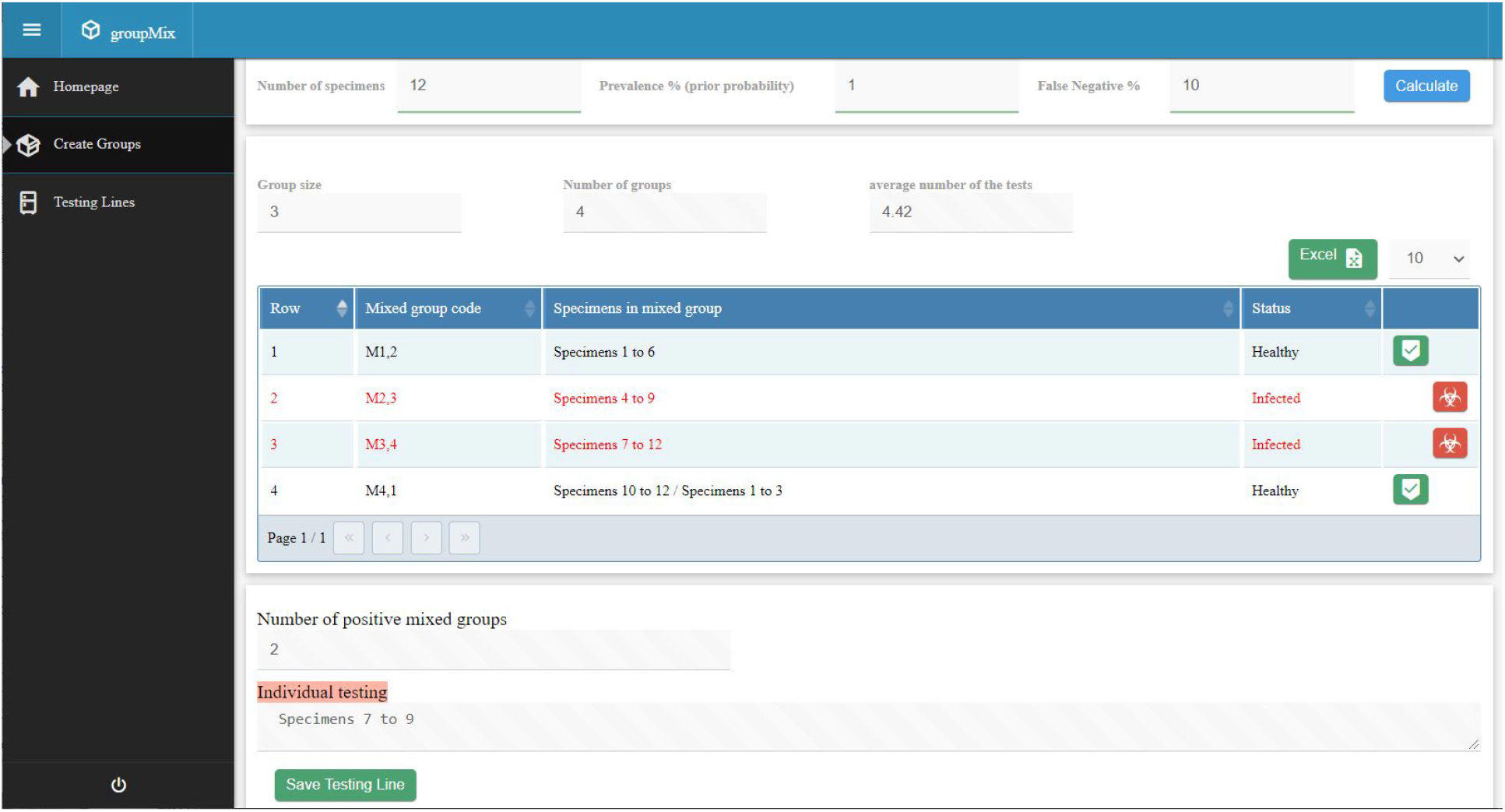
“Create Groups” section in the groupMix web interface for *N* = 12, *p* = 1%, FNR= 10%, and *n* = 3.

### B. Test Lines

In this section, we can see the list of saved test lines (Fig. B.3). Moreover, we can edit or delete each test line, and download the list of available test lines as an Excel spreadsheet.

**Fig. B.3.**
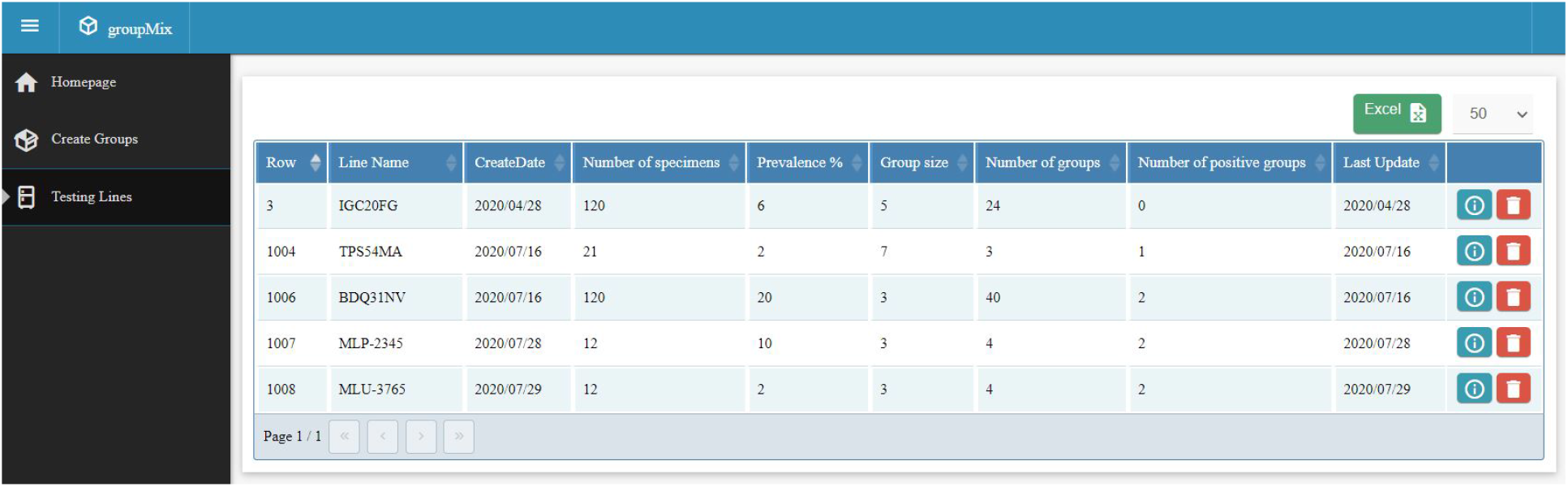
“Test lines” section in the groupMix web interface.

